# Socio-demographic correlate of knowledge and practice toward novel coronavirus among people living in Mosul-Iraq: A cross-sectional study

**DOI:** 10.1101/2020.09.11.20192542

**Authors:** Balsam Qubais Saeed, Rula Al-Shahrabi, Obasanjo Afolabi Bolarinwa

## Abstract

**Background:** Since the World Health Organization (WHO) announced that the 2019 novel coronavirus (2019-nCoV) is a worldwide pandemic, as the Iraqi authorities have started responding and taking action to control the spread of the pandemic. The knowledge and practices of the public play an important role in curbing the spreading of the virus by following the health guidelines. This study aimed to assess the socio-demographic correlate of knowledge and practices of Iraqi living in Mosul-Iraq towards novel coronavirus during its rapid rise.

**Methods:** A cross-sectional online survey of 909 participants was conducted among Mosul-Iraq between 20^th^ June to 1^st^ July 2020. The survey included three parts: 1) socio-demographic characteristics, 2) participants' knowledge, 3) participants' practices. T-test, ANOVA, chi-square, and binary logistic regression were used. A p-value less than 0.05 (p < 0.05) was considered statistically significant.

**Results:** The results showed knowledge and practice mean score of (12.91±1.67) and (21.56± 2.92) with cumulative knowledge and practice of 86% and 76% respectively towards 2019-nCoV. Socio-demographic characteristics such as age, marital status, gender, level of education and employment were statistically related with a higher mean score of knowledge and practice towards the virus as *P<0.05*.

**Conclusions:** We concluded that the majority of the respondents demonstrate a high level of knowledge and practices towards 2019-nCoV except respondents with socio-demographic characteristics such as those who were younger, male respondents, those with lower education and those unemployed as such campaigns that will increase the knowledge and encourage adequate preventive practice towards 2019-nCoV should be targeted towards this group.

## 1. INTRODUCTION

On 31^st^ December 2019, the novel coronavirus detected in Wuhan, China, on 11-March 2020, has been considered as a global pandemic by the (WHO), at the end of April, the virus has spread worldwide with fear-evoking death reports (1, 2).

The 2019 novel coronavirus (2019-nCoV) is highly contagious; it causes a respiratory illness that ranges from the common cold-like symptoms to more severe diseases (2). Most of the infected patients complaining from fever, shortness in breath, cough, loss of smell, and/or taste sensation and might be infected asymptomatically. In severe cases, patients might suffer from pneumonia, multiple organ dysfunction, and death. (3, 4).

The 2019-nCoV is a novel strain that is not specified in humans earlier, and it is a zoonotic illness that is transmitted between animals and people and may infect humans and animals. (5).

In Iraq, the first case of novel coronavirus was confirmed on 24^th^ -February-2020 in Al-Najaf city by people who have visited Iran. Therefore, at the end of March 13-2020, the Ministry of Health and Environment of Iraq declared that the total of confirmed cases of coronavirus is 101(6). The ministry of health (MOH) in Iraq started to respond and take action to control the infection, as well as the technical observation of (WHO), three governmental biological laboratories in Baghdad, Basrah, and Erbil, were opened for COVID-19 outbreak testing. (7). Initially, the measures that were taken by the Iraqi authorities nationwide have succeeded in managing to slow the spread of the virus, but the cases have risen sharply in the past two months. In the first week of August, Mosul city recorded thirty times increased in novel coronavirus cases reported compared to the previous months, while the reported national cases rose from 10,000 to 120,000 across the country (8).

Due to the escalating number of cases in Mosul city, many challenges were observed while trying to stop the virus from spreading, the limited number of tests that can be carried out per day, lack of quarantine facilities, medical instrument, hygienic preparation insufficiency, low hospital capacities, and no approved medicine or vaccine to prevent the 2019 n-CoV. Therefore, the preventive measures taken by the people are important to protect themselves and others from the virus infection and control the spread of the disease (9). Thus, managing this crisis hangs primarily on people’s knowledge and practices toward this virus and following all the precautions to prevent cross-infection and follow guidelines of the World Health Organization (WHO) and the Center for Disease Control and Prevention (CDC) (10, 11).

World Health Organization (WHO) declared precautional strategies to curb the spreading of infections. The none medical precautions are maintaining social distancing, avoid public gathering, avoid direct contact with infected people and use of personal protective equipment (PPE) like face masks. Also, personal hygiene recommendations such as hand-washing often with soap and water for at least 20 seconds, especially after touching surfaces, don’t touch the nose, eyes, and mouth with unwashed hands, and self-isolate when 2019-nCoV symptoms started (10).

The knowledge and practices of the public toward 2019 novel coronavirus play an important role in determining the willingness of Iraqi citizens to change their behavior and identify the kind of intervention that is needed to correct the misconceptions regarding the virus, highlight the poor knowledge toward the virus and disease, development of new preventive measures, develop COVID-19 awareness campaign, and take precautionary. Therefore, the current study aimed to assess the socio-demographic correlate of knowledge and practices of Iraqi in Mosul towards 2019 novel coronavirus during its rapid rise.

The results of this study are expected to provide baseline information about the level of knowledge and practices of Iraqi living in Mosul and highlight misperceptions related to preventive measures. The outcomes of the study will further hence better planning for effective awareness campaigns, required interventions and taking the appropriate action from local authorities.

## 1. MATERIAL AND METHODS

### Study design

A cross-sectional survey was adopted for the study using an online google form platform. The invitation of the respondents (Mosul–Iraqi) to participate in the survey was made on social media using the most popular media in Iraq, the social media messenger applications such as; WhatsApp and Facebook messengers were used in distributing the online google based questionnaire and participation was limited to Mosul–Iraqi 18 years and above.

### Study Settings

Mosul is located in Nineveh Governorate - Iraq, has a population of 3.5 million. Residents of Mosul city suffer from a fragile health system that barely meets their basic needs, as many health facilities were destroyed in 2017 (8).

### Survey and data collection

A total of 909 Mosul–Iraqi participated in our study between 20th June to 1st July 2020. The specified Sample size was by determining the lowest acceptable size of a demographic subgroup with a ±5% margin of error and a confidence level of 95% (12, 13). Giving to this, out of the total respondents of 1121 who filled the online google based questionnaire, only 909 were included. An incomplete survey of participants of 212 was excluded from the study, leaving us with a completion rate of 81%.

The survey was an adapted version of questionnaires published previously (14, 15,16). The survey was reviewed, and pilot tested by 21 Iraqi people and 3 faculty experts at the University of Mosul using WhatsApp and telephone interviews to correct any question then sent it to the target population. The survey consisted of an interface page and three main themes, with a total of 37 questions. The interface page included the title, objective of the study, information on participants’ privacy, and instructions to fill the survey. The three main themes included: 1) demographic information of participants such as gender, age, education level, marital status, employment status, coronavirus test, coronavirus result and Chronic diseases; 2) Knowledge related the 2019 novel coronavirus consisted of 15 questions divided to clinical presentations of virus (K1-K5), the spread of the virus (K6-K9) prevention (K10-K12) and the risk factors (K13-K15); 3) practices of participants toward novel coronavirus outbreak, which included 13 questions. The google based questionnaire was designed in Arabic to encourage adequate participation of the respondents since Arabic is the common language in Mosul city.

### Ethical approval

The Research Ethics Committee (RIC) at the University of Sharjah, UAE approved this study by the reference number is REC-20-05-31-01, as of 14/06/2020.

### Data and statistical analysis

Data analysis conducted using Statistical Package for Social Software (SPSS) version 22. Scale reliability was performed to ensure data consistency (Cronbach’s alpha coefficient = 0.729), indicating good consistency. The frequencies of demographic characteristics, knowledge, and practice answers along with descriptive statistics, were presented in mean ± SD, while qualitative data were presented in frequency (number\percent). Participants’ knowledge and practice scores were compared with demographics factors using independent-samples t-test, one-way analysis of variance (ANOVA).

To measure the knowledge, participants were given “yes,” “no,” and “not sure” answer options to each survey question. A true answer to each question was marked with 1 score, while false answers and not sure were marked with 0 scores. Scores of total knowledge ranged from 0-15; a higher score signals a better level of knowledge. To practice measures, participants were given "always", "sometimes", and " never” answer options to each item; the always option was marked for 2 scores, while sometimes was for 1 score, and rarely was for 0 scores. The total practice scores ranged from 0-26. The lowest and highest score of participants' knowledge was 7 and 15, respectively, while the lowest and highest score of practice was 14 and 26, respectively.

Pearson’s chi-square was used to determine the association between the explanatory and outcome variables. We examined the factors associated with good knowledge & practice and poor knowledge & practices by using binary logistic regression analyses. A p-value of less than 0.05 (p < 0.05) was considered statistically significant.

To identify the factors that were significantly associated with good or poor knowledge and practices, a mean knowledge score of more than 12 indicated as good knowledge, while less than 12 assigned as poor. Similarly, a mean practice score above 21 indicated as good practice and a mean practice score below 21 as poor practice. Factors were selected with a backward stepwise method, and the reference category was selected based on the higher total mean. Unstandardized regression coefficients (β) and odds ratios (ORs) and their 95% confidence intervals (CIs) were used to quantify the associations between cofactors with knowledge and practice.

## 2. RESULTS

### Socio-demographic characteristics

The results showed that the majority of 61.4% were females, and 38.6% were males. More than half of the participants, 54.5%, aged 30-49 years. Around 60.5% of respondents were married, while 31.7% and 7.9% were single, and others (divorced and widows), respectively. About 62.4% were holding a bachelor’s degree, while 25.7, 11.9% were holding postgraduates and diploma or below, respectively. Moreover, almost 64.5% were employed, while a smaller number of participants, 21.8%, 13.9%, were unemployed and students, respectively. Some of the participants, 7.9%, have been tested for COVID-19, while 92.1% didn’t do the COVID-19 est. However, 1.5% of tested respondents were positive, and 98.5% of the participants have been negative tests. Figure 1 summarized the frequency of respondents according to demographic characteristics.

**Fig. 1.**
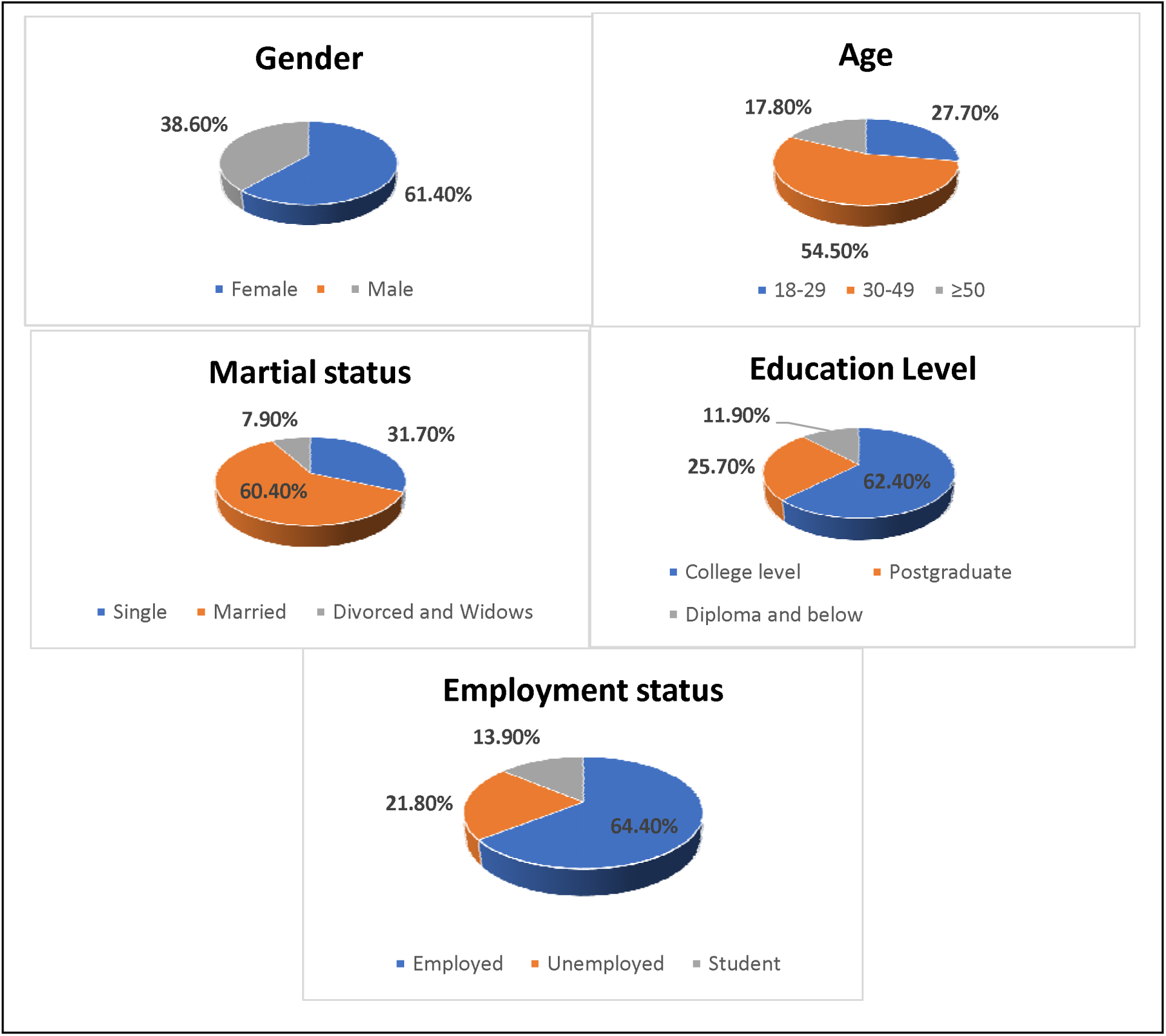
Socio-demographic characteristics of the participants Mosul, Iraq (n = 909).

### Prevalence of chronic diseases reported by Iraqi participants

In our study, 82.4% of the participants were healthy, while 17.6% had chronic diseases. The most common chronic diseases were diabetes 6%, followed by asthma at 3.30%. Moreover, 2.75%, 2.40%, 1.54%, 1.54%, 1.20% had chronic kidney disease, severe obesity, heart conditions, chronic liver disease, and chronic lung diseases, respectively, as shown in Figure 2.

**Figure. 2.:**
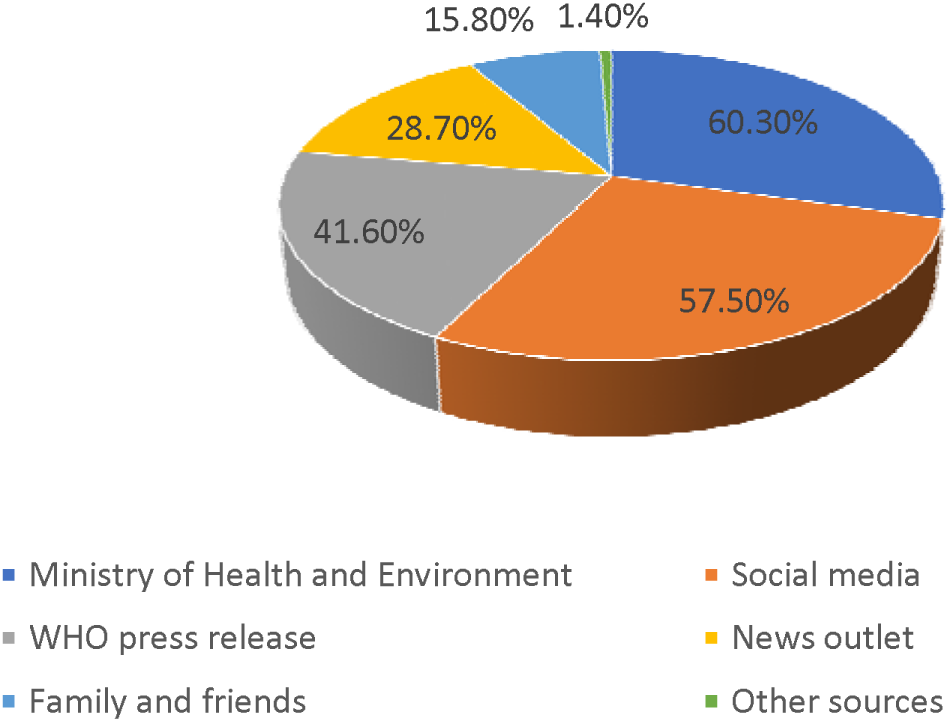
Prevalence of chronic diseases reported by people in the Mosul, Iraq (n = 909)

### Knowledge towards novel coronavirus

The mean knowledge score (±SD) of 15 questions was (12.91±1.67). The correct percentage rate was 86.08%. Most of the participants, 92.1%, answered correctly that novel coronavirus caused by a virus; the majority, 94.1%, knew that the incubation period range of this virus is between 2-14 days. Almost all participants had a high knowledge of 96% about the symptoms of a novel coronavirus; about 72.3% of respondents knew that no vaccine toward this virus.

Moreover, 63.4% knew that no treatment was approved toward novel coronavirus until now. When we asked if this virus is spread via respiratory droplets of infected people, 93.1% answered correctly.

Similarly, A high proportion of 93.1% of the participants agreed that the virus could be transmitted via touching contaminated surfaces. Also, 82.2% indicated that this virus was transmitted through the eyes, nose, and mouth. Just over half, 59.4%, reported that the infected person having no fever could infect healthy people and the majority of participants, 94.1% recorded that children and young adults have to take measures toward novel coronavirus.

About 76.2% of respondents reported the individuals should stay at home and go out only when necessary, while all of the participants 100% agreed that they should avoid going to crowded places, and the infected person with this virus should be immediately isolated in a proper place. Most of the respondents, 96%, answered that the virus is more dangerous for those with chronic disease patients and the elderly, while 79.2% believed that smokers are more vulnerable to this virus. The knowledge of participants toward novel coronavirus is displayed in Table 1.

**Table 1.** General knowledge of participants about novel coronavirus in Mosul, Iraq (n= 909)

| Knowledge questions |  | Frequency (%) | MEAN±SD |
| --- | --- | --- | --- |
| K:1 Novel coronavirus is caused by virus | Yes | 837 (92.1) | 1.13 ±0.460 |
|  | No | 27 (3.0) |  |
|  | Not sure | 45 (5.0) |  |
| K:2 Incubation period range of novel coronavirus is 2-14 day | Yes | 855 (94.1) | 1.07 ±0.291 |
|  | No | 45 (5.0) |  |
|  | Not sure | 9 (1.0) |  |
| K3: The main clinical symptoms of Novel coronavirus are fever, dry cough, tiredness, and breathing difficulties | Yes | 873 (96.0) | 1.08 ± 0.390 |
|  | No | 0(0) |  |
|  | Not sure | 36 (4.0) |  |
| K4: No vaccine against novel coronavirus | Yes | 657 (72.3) | 2.10 ±0.517 |
|  | No | 81 (8.9) |  |
|  | Not sure | 171 (18.8) |  |
| K5: No active treatment for novel coronavirus | Yes | 576 (63.4) | 2.23 ±0.561 |
|  | No | 63 (6.9) |  |
|  | Not sure | 270 (29.7) |  |
| K6: Novel coronavirus spreads via respiratory droplets of infected people | Yes | 846 (93.1) | 1.13 ±0.481 |
|  | No | 9 (1) |  |
|  | Not sure | 54 (5.9) |  |
| K7: Novel coronavirus is spread through touching contaminated surfaces | **Yes** | **846 (93.1)** | 1.13 ±0.481 |
|  | No | 9 (1) |  |
|  | Not sure | 54 (5.9) |  |
| K8: Novel coronavirus can be transmitted through eyes, in addition to nose and mouth | **Yes** | **747 (82.2)** | 1.24 ±0.548 |
|  | No | 108 (11.9) |  |
|  | Not sure | 54 (5.9) |  |
| K9: A person with novel coronavirus having no fever cannot infect others | **Yes** | **162 (17.8)** | 2.05 ±0.636 |
|  | No | 540 (59.4) |  |
|  | Not sure | 207 (22.8) |  |
| K10: Children and young adults don't need to take measures to prevent the infection by the novel coronavirus | **Yes** | **18 (2)** | 2.02 ±0.243 |
|  | No | 855 (94.1) |  |
|  | Not sure | 36 (4) |  |
| K11: We should stay at home and go out only when it is necessary | **Yes** | **693 (76.2)** | 1.48 ±0.852 |
|  | No | 216 (23.8) |  |
|  | Not sure | 0 (0) |  |
| K12: To prevent the spread of novel coronavirus, individuals should avoid going to crowded places if it's not necessary | **Yes** | **909 (100)** | 1.00 ±0.00 |
|  | No | 0(0) |  |
|  | Not sure | 0(0) |  |
| K13: The novel coronavirus may be more dangerous in patients with chronic diseases and elderly | **Yes** | **873 (96)** | 1.07 ±0.352 |
|  | No | 9 (1) |  |
|  | Not sure | 27 (3) |  |
| K14: People who have contact with someone infected with the novel coronavirus should be immediately isolated in a proper place | **Yes** | **909 (100)** | 1.00 ±0.00 |
|  | No | 0(0) |  |
|  | Not sure | 0(0) |  |
| K15: Smokers are likely to be more vulnerable to novel coronavirus | **Yes** | **720 (79.2)** | 1.31 ±0.641 |
|  | No | 99 (10.9) |  |
|  | Not sure | 90 (9.9) |  |
| **Total knowledge level** |  | **86.08%** |  |

### Practice towards novel coronavirus

The mean practice score (±SD) of 13 questions was (21.56± 2.92). The correct percentage rate was 75.8%. We found that 92.1% of the participants started washing their hands frequently during a novel coronavirus period. Similarly, 92.1% indicated they were used sanitizer if the soap is not available, while usual handwashing with soap for 20 seconds was recorded by 77.2% of respondents. Almost 83.2% of participants wearied a mask when they go outside the home. Three-quarters, 72.3% of respondents maintain space between them and others when going outside, and only 36.6% maintain the 2-meter distance between them and others to prevent transmission of the virus.

About 88% of the Iraqi respondents stopped going to crowded places recently. However, two-thirds of the respondents, 69.3% and 65.3%, reported that they stopped visiting and kissing their relatives or friends when meeting them, while 77.2% stopped the handshake during the greeting with others. Besides, 81.2% were aware of the essential of sanitizing their hands after using cash, and 75.2% of participants were aware of the importance of avoiding sharing their food with others. The participant’s practices toward novel coronavirus prevention presented in Table 2.

**Table 2.**
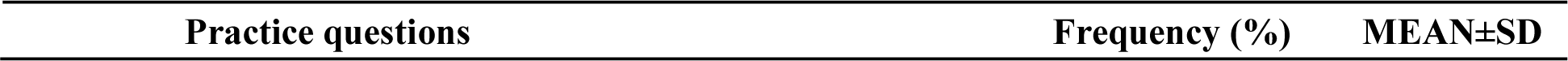
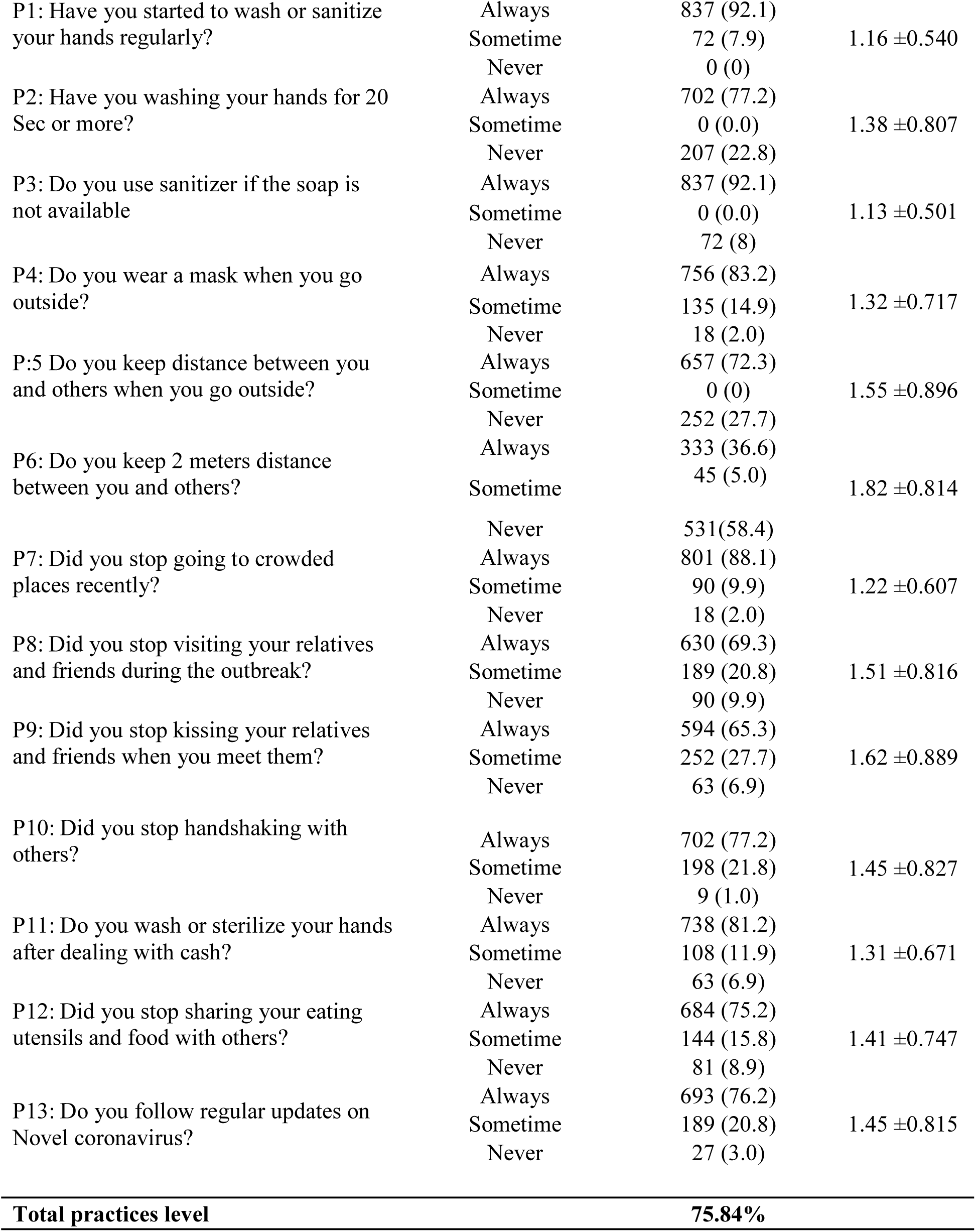
Practices of participants toward novel coronavirus in Mosul, Iraq (n= 909)

| Practice questions | Frequency (%) | MEAN±SD |
| --- | --- | --- |

|  |  |  |  |
| --- | --- | --- | --- |
| P1: Have you started to wash or sanitize your hands regularly? | Always | 837 (92.1) | 1.16 ±0.540 |
|  | Sometime | 72 (7.9) |  |
|  | Never | 0 (0) |  |
| P2: Have you washing your hands for 20 Sec or more? | Always | 702 (77.2) | 1.38 ±0.807 |
|  | Sometime | 0 (0.0) |  |
|  | Never | 207 (22.8) |  |
| P3: Do you use sanitizer if the soap is not available | Always | 837 (92.1) | 1.13 ±0.501 |
|  | Sometime | 0 (0.0) |  |
|  | Never | 72 (8) |  |
| P4: Do you wear a mask when you go outside? | Always | 756 (83.2) | 1.32 ±0.717 |
|  | Sometime | 135 (14.9) |  |
|  | Never | 18 (2.0) |  |
| P5: Do you keep distance between you and others when you go outside? | Always | 657 (72.3) | 1.55 ±0.896 |
|  | Sometime | 0 (0) |  |
|  | Never | 252 (27.7) |  |
| P6: Do you keep 2 meters distance between you and others? | Always | 333 (36.6) | 1.82 ±0.814 |
|  | Sometime | 45 (5.0) |  |
|  | Never | 531 (58.4) |  |
| P7: Did you stop going to crowded places recently? | Always | 801 (88.1) | 1.22 ±0.607 |
|  | Sometime | 90 (9.9) |  |
|  | Never | 18 (2.0) |  |
| P8: Did you stop visiting your relatives and friends during the outbreak? | Always | 630 (69.3) | 1.51 ±0.816 |
|  | Sometime | 189 (20.8) |  |
|  | Never | 90 (9.9) |  |
| P9: Did you stop kissing your relatives and friends when you meet them? | Always | 594 (65.3) | 1.62 ±0.889 |
|  | Sometime | 252 (27.7) |  |
|  | Never | 63 (6.9) |  |
| P10: Did you stop handshaking with others? | Always | 702 (77.2) | 1.45 ±0.827 |
|  | Sometime | 198 (21.8) |  |
|  | Never | 9 (1.0) |  |
| P11: Do you wash or sterilize your hands after dealing with cash? | Always | 738 (81.2) | 1.31 ±0.671 |
|  | Sometime | 108 (11.9) |  |
|  | Never | 63 (6.9) |  |
| P12: Did you stop sharing your eating utensils and food with others? | Always | 684 (75.2) | 1.41 ±0.747 |
|  | Sometime | 144 (15.8) |  |
|  | Never | 81 (8.9) |  |
| P13: Do you follow regular updates on Novel coronavirus? | Always | 693 (76.2) | 1.45 ±0.815 |
|  | Sometime | 189 (20.8) |  |
|  | Never | 27 (3.0) |  |
| **Total practices level** |  | **75.84%** |  |

### Sources of information on COVID-19

Participants indicated that the Ministry of Health and Environment in Iraq (MOH) and social media such as Twitter, Facebook, YouTube, WhatsApp, Instagram, and Snapchat were the main sources of information about novel coronavirus pandemic with 60.3% and 57.5% respectively, followed by World Health Organization (WHO) press release 41.6%, while 28.7%, 15.8%, and 1.4% reported that they received their information from the news outlet, family and friends, and other sources, respectively as presented in Figure 3.

### Level of knowledge (K) and practice (P) as per socio-demographic characteristics of participants

The knowledge level score (out of 15) showed a significant association across socio-demographic characteristics such as gender, age, education levels, marital status, and employment status (p <0.005). The practice level score (out of 26) also showed a significant association between gender and age-groups (p <0.005) while there was no significant association between marital status ((p=0.061), educational (p=0. 385) and employment (p=0.084) with the practice of the participants.

The results indicated that females had a higher mean score of knowledge (13.19±1.70) and practice (21.85±2.61) than males, aged group of participants above 50-years-old having the highest score of knowledge (14.11±0.87) and practice (22.50±2.32) compared with other age groups. Moreover, widows and divorced women's knowledge (13.37±1.32) were higher than singles and married participants; however, there were no significant differences in practice. The mean score of knowledge (13.26±1.51) and practice (21.75 ±2.77) of participants with high education degrees were better than participants with lower educational degrees. Employed respondents showed a higher-level score of knowledge (13.12± 1.61) than non-employed and students’ participants. While there were no significant differences in employment status in practice, neither education levels nor employment status had any significant differences in practice, as depicted in table 3.

**Table 3.** Association between socio-demographic characteristics with knowledge (K) and practices (P) of participants.

| Knowledge of participants |  |  |  |  |  |
| --- | --- | --- | --- | --- | --- |
| Variables |  | Frequency (%) | MEAN±SD | t/F | P-value |
| Gender | Female | 558 (61.4%) | 13.19±1.70 | 6.57 | 0.000 |
|  | Male | 351 (38.6%) | 12.46±1.51 |  |  |
| Age | 18-29 | 252 (27.7%) | 12.35±1.63 | 64.54 | 0.000 |
|  | 30-49 | 495 (54.5%) | 12.80±1.70 |  |  |
|  | ≥ 50 | 162 (17.8%) | 14.11±0.87 |  |  |
| Marital status | Single | 288 (31.7%) | 12.40±1.47 | 20.83 | 0.000 |
|  | Married | 549 (60.4%) | 13.11±1.74 |  |  |
|  | Others | 72 (7.9%) | 13.37±1.32 |  |  |
| Education level | College degree | 567 (62.4%) | 12.84±1.62 | 9.28 | 0.000 |
|  | Postgraduate degree | 234 (25.7%) | 13.26±1.51 |  |  |
|  | Others | 108(11.9%) | 12.50±2.07 |  |  |
| Employment status | Employed | 585 (64.4%) | 13.12±1.61 | 17.44 | 0.000 |
|  | Unemployed | 198 (21.8%) | 12.72±1.93 |  |  |
|  | Student | 126 (13.9%) | 12.21±1.21 |  |  |

| Practices of participants |  |  |  |  |  |
| --- | --- | --- | --- | --- | --- |
| Variables |  | Frequency (%) | MEAN±SD | t/F | P-value |
| Gender | Female | 558 (61.4%) | 21.85±2.61 | 3.8 | 0.000 |
|  | Male | 351 (38.6%) | 21.1±3.31 |  |  |
| Age | 18-29 | 252 (27.7%) | 21.64 ±2.89 | 12.14 | 0.000 |
|  | 30-49 | 495 (54.5%) | 21.21±3.04 |  |  |
|  | ≥ 50 | 162 (17.8%) | 22.50±2.32 |  |  |
| Marital status | Single | 288 (31.7%) | 21.87 ±2.87 | 2.80 | 0.061 |
|  | Married | 549 (60.4%) | 21.45 ±2.99 |  |  |
|  | Others | 72 (7.9%) | 21.12 ±2.48 |  |  |
| Education level | College degree | 567 (62.4%) | 21.4 ± 3.07 | 0.955 | 0.385 |
|  | Postgraduate degree | 234 (25.7%) | 21.75 ±2.77 |  |  |
|  | Others | 108(11.9%) | 21.73 ±2.36 |  |  |
| Employment status | Employed | 585 (64.4%) | 21.63±2.96 | 2.48 | 0.084 |
|  | Unemployed | 198 (21.8%) | 21.18±2.92 |  |  |
|  | Student | 126 (13.9%) | 21.85±2.67 |  |  |

### Binary logistic regression analysis

Table 4 shows the binary logistic regression analysis on variables significantly correlated with knowledge and practice (good and poor) about 2019 novel coronavirus.

**Table 4:**
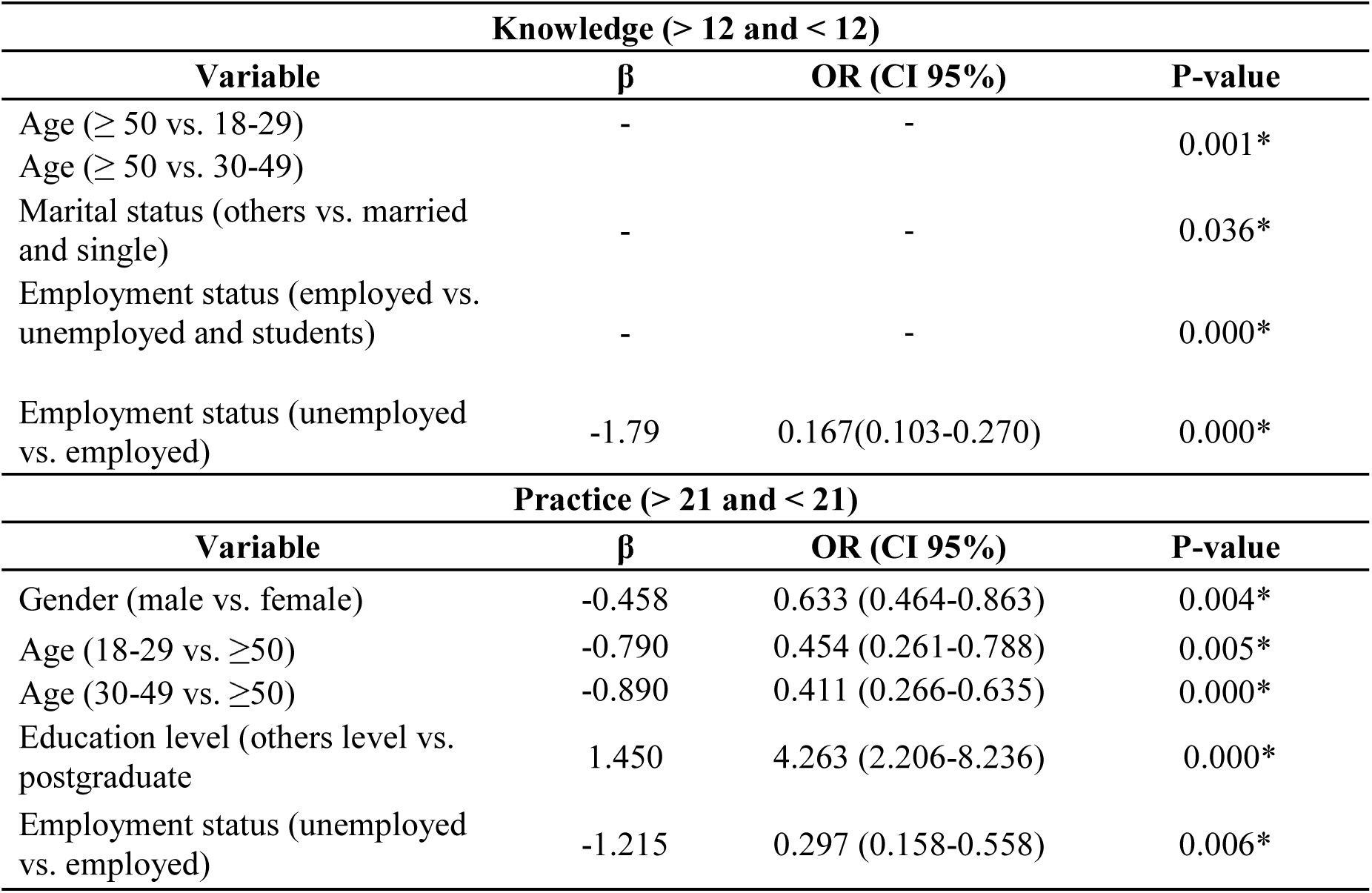
Binary logistic regression analysis on factors significantly associated with mean knowledge and practices about novel coronavirus of the participants, Mosul-Iraq

The Odds ratios (ORs) and their 95% confidence intervals (CIs) in a bid to quantify the relationship between socio-demographic characteristics and the knowledge score (>12 and <12), and between socio-demographic characteristics and the practices score (>21 and < 21). Overall, the analysis presents a significant relationship between knowledge with age, marital status, and employment status. The same table shows that the age of ≥50 vs other ages reported better knowledge, divorced & widows vs. married and single participants were significantly related with a higher mean score, employed people were significantly related with the better knowledge vs unemployed and retired respondents (β:-1.790, OR: 0.167, CI: 0.103-.270). Moreover, gender, age, education and employment status were significantly associated with good practices. Female gender vs male (β: -0.458 OR: 0.633, CI 0.464-0.863), age group of ≥50 had higher practice score than the youngest group of 18-29 and 30-49 (β: -0.790, OR: 0.454, CI: 0.261-0.788), (β: -0.890 OR: 0.411, CI: 0.266-0.635), respectively. However, participants with a college-level degree and below showed a better practice (β 1.450 OR: 4.263, 95% CI: 2.206-8.236) than participants with a postgraduate degree, while, employment statues of employed reported a better score than unemployed participants to (β: -1.215, OR: 0.297, CI: (0.158-0.558), as indicated in Table 4.

## 3. DISCUSSION

The knowledge and practices of the public play an important role in prevention by following the health guidelines to control the spread of 2019-nCoV. The knowledge and practices of the general population about novel coronavirus have changed during the pandemic as a defense line against the disease.

The provided baseline data regarding the level of individuals' knowledge on clinical presentations, transmission, prevention, and risk factors of novel coronavirus virus will help highlight malpractices related to preventive measures hence making it critical for local authorities to plan suitable strategies to prepare and manage the spread of the virus.

The current study resulted that the participants' knowledge was high (86%) and had good measures of practice toward 2019 novel coronavirus (76%). Similar to our findings, several studies done in many countries have reported high levels of knowledge about novel coronavirus, among the general population in Malaysia 80.5%, Chinese residents 90%, Saudi Arabia population 81.5%, and among healthcare workers in Pakistan 93.2%. (14, 4, 15, 16).

The high level of knowledge among the public may be due to most of the participants having a college degree or higher, or due to the high level of media coverage, including all media outlets and the impact of the pandemic on social life mandating that people follow.

The current results showed that most of the participants depend more on the ministry of health and social media to get their information about the 2019 novel coronavirus, In contrast to other studies among Jordanian, Egyptian and Pakistani populations that were using mostly social media as the main source of information (17, 18, 19).

The study found that the majority of respondents had a good level in prevention and control measures toward novel coronavirus, indicating that the practices of some respondents were very good toward novel coronavirus. That the results reported on practice toward 2019-nCoV among the respondents were similar to those reported in the Malaysian population (14) while being less than the level of practices among Chinese residents (4).

This level of practice among the respondents attributed that the Iraqi government didn't take drastic measures in reducing the spread of the disease, in addition to the low number of cases in Mosul at the beginning of the pandemic (20).

Our results using t-tests, ANOVA and logistic regression analysis showed that there was better knowledge and accurate practices associated significantly with female gender, respondents above 50 years old, employed respondents, higher education, and married respondents.

Females and mothers are expected to show better knowledge and practices towards the 2019 novel coronavirus precautions and preventions. Similar studies in Malaysian and Saudi Arabia indicated that females had more knowledge regarding novel coronavirus than males (14, 15).

The high level of knowledge and practice among the participants that are above the age of 50 in our study is possibly due to understanding the higher risk of contraction and complications of the disease on the elderly and people with chronic diseases (21). In our study, the respondents that hold a higher degree level had a greater level of knowledge than the others. Similar findings were reported within Malaysian and Pakistani university populations (14,19). A great majority of respondents confirmed that the novel coronavirus disease is more dangerous in patients with chronic diseases and the elderly. This has been confirmed from many studies published regarding the 2019 novel coronavirus disease in China (22, 23).

Our results reported that 17.5 % of the respondents had chronic diseases such as diabetes, asthma, and severe obesity were the common diseases among our participants. A similar study among Iraqi adults indicated that the common non-communicable diseases were hypertension (13.3%) and overweight or obese (54.6%). Another study showed that diabetes and hypertension were the most prevalent diseases among Iraqi people (24, 25).

Our results were compatible with many studies that showed similar significance in terms of better knowledge and practice among the educated and employed people. (4, 14, 15,26, 27).

Finally, the results indicated that more informative novel coronavirus efforts and more intense health education should be directed toward respondents with the following socio0demographic characteristics; male respondents, respondents with lower educational levels, younger respondents, and unmarried respondents.

### Limitations

Since the study is a cross-sectional study, it was conducted within a short time during the pandemic. Moreover, this study was an online survey that expected that the people with a higher level of education would respond to the survey; as such, it doesn’t give privilege to the uneducated population and those with limited access to the internet.

## 4. CONCLUSIONS

In general, the current study provided a comprehensive screening of the knowledge and practices of the population in Mosul city toward the 2019 novel coronavirus. The participants had a high level of knowledge about the virus, and good practice towards using protective measures, which is significant towards controlling the spread of the virus. The study recommends developing informative novel coronavirus related campaigns targeted specifically towards younger males, lower educated, and unemployed people were living in Mosul city. Furthermore, interventions that encourage the provision and use of free personal protective equipment (PPE) such as face mask should be developed as this will help curb the spreading of the virus. In the same vein, the Iraqi government should take more effort to control the spread of the disease.

## Data Availability

All data referred included in the manuscript

## ACKNOWLEDGMENT

The authors thank the University of Sharjah and respondents who agree to participate in the study. Also, we gratefully acknowledge Mr. Ahmed Omar Address - College of Medicine for the critical edition of English grammar.

## AUTHOR CONTRIBUTIONS

B.Q.S. Conceptualization, design of the study, data collection, resources, and wrote the paper, R.A contributed to the statistical analysis, and O.A.B Revise and edit the text, All authors have read and agreed to the published version of the manuscript.

